# A Pre-Post Single-Arm Trial of a Multicomponent Intervention for Self-Efficacy Enhancement among an Online Support Group of Primary Caregivers of Children with Neuro Muscular Disorders: SEE-NMD Protocol

**DOI:** 10.64898/2026.05.04.26352405

**Authors:** Nashit Irfan Aziz, Wardah Khalid, Sara Khan

**Author notes:** These authors contributed equally to this work (Nashit Irfan Aziz, Wardah Khalid).

## Abstract

**Introduction:** Spinal Muscular Atrophy (SMA) and Duchenne Muscular Dystrophy (DMD) are progressive genetic neuromuscular disorders imposing substantial caregiving demands on families. In Pakistan, where disease-modifying therapies remain largely inaccessible, care is predominantly supportive, frequently resulting in psychological distress and diminished caregiver confidence. An individual’s belief in their capacity to execute required behaviours, self-efficacy, is a recognised determinant of caregiving quality, yet evidence on caregiver self-efficacy in SMA and DMD within low-resource settings remains sparse. This study aims to evaluate the effect of a multicomponent, digitally delivered intervention on self-efficacy among primary caregivers of children with SMA and DMD in Pakistan.

**Methods and Analysis:** This single-group, pre-post study will recruit 30 primary caregivers of children with SMA or DMD enrolled in the Treat-NMD Registry of Pakistan. An eight-week intervention will be delivered via online support groups, comprising educational video clips, expert-led live sessions, pictorial guides, and progressively tapering audio reminders. The primary outcome, self-efficacy, will be measured using a culturally adapted, content-validated Urdu version of the DMD Caregiver Self-Efficacy Scale (DMD-CSES), assessed at baseline (T0) and eight weeks post-intervention (T1). Pre-post scores will be compared using a paired t-test or Wilcoxon signed-rank test depending on data distribution, with analyses conducted in STATA Version 17.

**Ethics and Dissemination:** Ethical approval has been granted by the Ethics Review Committee of Aga Khan University (2025-11875-37040). Verbal informed consent will be obtained from all participants, with confidentiality maintained throughout. Findings will be disseminated via peer-reviewed publication, conference presentations, and shared with the Treat-NMD registry network.

**Trial registration number:** <u>NCT07356063</u>, Date of registration: 11/January/2026

## Introduction

Spinal Muscular Atrophy (SMA) and Duchenne Muscular Dystrophy (DMD) are progressive, genetic neuromuscular disorders that substantially affect children and their families. SMA is characterised by degeneration of motor neurons in the spinal cord, resulting in progressive limb and respiratory muscle weakness (1), while DMD is an X-linked recessive disorder marked by progressive muscle degeneration and weakness (2).

The prevalence of SMA is approximately 1–2 per 100,000 persons, and its incidence is estimated at around 1 in 10,000 live births (3). However, these pooled estimates are largely derived from older studies with small sample sizes and limited geographical scope, with an overrepresentation of European populations. Similarly, the burden of DMD is often estimated using databases and registries. A comprehensive systematic review published in 2020 reported a pooled global prevalence of 7.1 (95% CI: 5.0–10.1) and a birth prevalence of 19.8 (95% CI: 16.6–23.6) per 100,000 live births (4). In Pakistan, Bisma et. al analysed 215 SMA patients from the country’s inaugural registry (MDR-PK) (5) and 130 DMD patients (6) from 2019–2024, suggesting that both conditions are present; however, neither study provides population-level incidence or prevalence estimates.

Disease modifying drugs are now available for both neuromuscular disorders, however in Pakistan their use is limited due to availability and financial constraints. The main stay of treatment remains supportive management. This requires specialised and time-intensive caregiving, often extending over several years and increasing in complexity as the child’s condition progresses. Disease progression frequently results in worsening functional decline, discomfort, and respiratory dysfunction (7), intensifying the need for continuous monitoring, medication administration, physical support, and use of assistive devices. These demands may contribute to social isolation, financial strain, and psychological distress among caregivers, adversely affecting their overall quality of life and diminishing confidence in their ability to manage caregiving responsibilities (8). Thus, understanding this confidence level or ‘self-efficacy’ is of critical importance. Self-efficacy is defined as an individual’s belief in their capacity to execute behaviours required to achieve desired outcomes (9).

In the context of chronic childhood illnesses, caregiver self-efficacy plays a pivotal role in sustaining long-term care and adapting to evolving disease-related challenges. Caregivers with higher self-efficacy are more likely to approach caregiving tasks with confidence, persist despite setbacks, and report lower levels of stress and burnout (10). Conversely, lower self-efficacy can result in feelings of helplessness, anxiety, and depression, potentially compromising both: caregiver’s well being and child’s quality of care. Despite caregiver burden, evidence investigating a caregiver’s self-efficacy for these neuromuscular conditions remains limited. A study validated a scale on the self efficacy of caregivers of DMD patients (11), meanwhile another reported low self-efficacy among parents of children with SMA, with a mean score of 46.18 ± 9.26 on the General Self-Efficacy Scale (GSES), where observed scores ranged from 33 to 73 (12).

Support groups may offer caregivers opportunities to share lived experiences, enhance disease-related knowledge, and access emotional support from individuals facing similar challenges (13). However, the concept of such is also in its infancy in Pakistan, with no formal organisations targeting neuromuscular disorders. To add further, conventional in-person support groups may be even more inaccessible here due to geographical barriers, time constraints, or transportation difficulties. Online (internet-based) support groups have therefore emerged as a practical alternative in literature (14), as it offers better flexibility of time, improved accessibility, and a degree of anonymity that may be particularly beneficial for caregivers managing demanding responsibilities. These platforms can facilitate peer-to-peer interaction, professional guidance where available, and access to relevant educational resources from within their home environment.

Evidence from caregiver-focused interventions in neurodevelopmental disabilities suggests that structured parent training programmes can significantly improve parental self-efficacy, with a meta-analysis reporting a standardised mean difference of 0.60 (95% CI: 0.38–0.83) relative to baseline (15). In addition, a meta-analysis of internet-based group support interventions for family caregivers across diverse chronic conditions reported statistically significant improvements in caregiver social support and self-efficacy, suggesting that online peer and professional support may enhance caregiver confidence (16). These findings suggest potential applicability to caregivers of children with SMA and DMD, although condition-specific evidence remains limited.

Health systems in low and middle-income countries (LMICs) such as Pakistan are often oriented toward acute care and lack the infrastructure, trained workforce, and resources necessary for comprehensive management of chronic conditions, including SMA/DMD. Redesigning chronic care in LMICs requires attention to quality communication, and sustained engagement beyond episodic treatments, yet these elements remain underutilised (17).

In this context, a multicomponent intervention delivered through an online support group may provide a feasible and scalable strategy to address the informational, emotional, and behavioural needs of caregivers. However, at present, structured interventions specifically designed for SMA/DMD caregivers remain limited, both globally and in Pakistan.

Therefore, this study aims to evaluate the effect of a multicomponent intervention on self-efficacy among primary caregivers of children with SMA or DMD. By generating evidence on caregiver self-efficacy and the potential impact of structured online support, this study seeks to inform future interventions and provide evidence-based recommendations for healthcare providers, support organisations, and policymakers involved in the care of children with neuromuscular disorders.

## Materials and Methods

### Study Design and Setting

This study will be a single-arm pre–post interventional study conducted online on caregivers of patients enrolled in the Treat-NMD Registry of Pakistan (also called the Muscular Dystrophy Registry of Pakistan (18). Treat-NMD Registry of Pakistan is the national subdivision of Treat-NMD Global Registry Network (a federated network of 67 independent patient registries that collects high-quality information on rare neuromuscular dystrophy (NMD) patients worldwide (19). In Pakistan, the method of collecting data for registry involves contacting caregivers of SMA/DMD patients via phone calls, since majority of the patients are children.

All study participants will be administered the study scale at two time points: T0 (baseline), and T1 (8 weeks after intervention), to assess change in study’s outcome (self-efficacy), as demonstrated in (**Fig 1.)**

**Fig 1.**
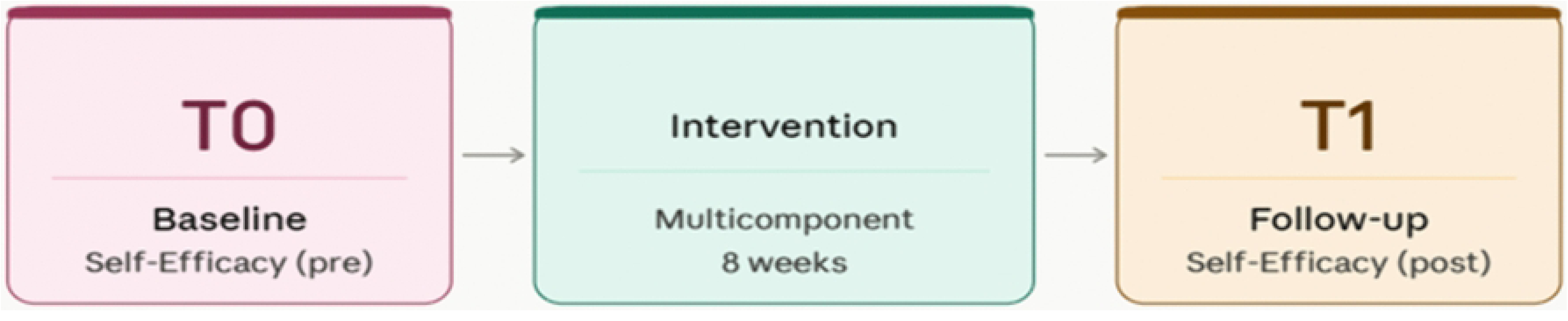
The study’s summary.

The reporting of this protocol follows the SPIRIT 2025 Statement (20) (S1 Checklist) for interventional trials and the TIDieR checklist (21) (S2 Checklist) for intervention description. Completed checklists for both are provided as supplementary files (S1 Checklist and S2 Checklist, respectively).

### Sample size

Sample size calculation was performed using NCSS 2024 (NCSS LLC, Kaysville, Utah, USA), based on a one-tailed t-test with a significance level of 0.05 and desired power of 80%. A one-tailed test was used as we expect consistent improvement in self-efficacy with no potential harms. Since there are no prior studies of this intervention in Pakistani SMA/DMD caregivers, we conservatively assumed a medium effect size (Cohen’s d = 0.5). This corresponds to an expected mean change of 0.5 points on a 5-point item scale (with an assumed SD of 1.0), which represents a moderate and clinically meaningful improvement (22). Based on these parameters, a sample size of 27 was obtained. After adjusting for 10% anticipated dropout rate, the sample size was 30. However, we acknowledge that this sample size estimate is preliminary and based on limited data. The data collected during this study can be used to calculate a more accurate effect size, which will inform future sample size calculations for larger-scale trials (23). This approach aligns with recommendations for iterative sample size refinement in the absence of strong prior evidence (24).

### Participants

These will be the primary caregivers. Primary caregivers are the individuals (from family) who provide most of the care and support for the child with SMA or DMD (11). The eligibility criteria for this study will be as follows:

Inclusion:

1. Primary caregiver of a child diagnosed with SMA or DMD.
2. Access to a smartphone with internet connectivity.

Exclusion:

1. Caregivers who are unable to understand the language of communication (Urdu).
2. Caregivers whose children have had their diagnosis for less than one year.
3. Caregivers who are currently participating in another support group intervention.

### Recruitment

Participants will be recruited using a non-probability, purposive sampling approach from the Treat-NMD Pakistan registry. Eligible participants will include a minimum of one year’s caregiving experience, at the time of recruitment. This threshold ensures participants have transitioned past the initial ‘turbulent’ phase of diagnosis into a more established caregiving role. This approach is supported by evidence indicating that caregiver burden often stabilizes after the first year; for instance, Klietz et al. (25) observed no significant fluctuations in Parkinson’s caregiver burden between the one-year mark and subsequent follow-ups. Furthermore, recent reviews identify a ‘stable average trajectory’ of burden as the most prevalent long-term pattern (26), suggesting that experience beyond 12 months provides a more consistent baseline for intervention.

Recruitment will be conducted between May and June 2026 by a trained recruiter (research associate or study investigator). Caregivers listed in the Treat-NMD registry, with specific characteristics, will be approached for participation, and recruitment will be conducted sequentially on the year of diagnosis, beginning with earlier records and progressing to more recent entries.

Contact will be made via telephone. Up to three contact attempts, spaced over time, will be made for each caregiver. Individuals who do not respond after three attempts will be classified as non-responders and will not be pursued further unless they initiate contact. Eligibility screening will be conducted during the call using predefined criteria. Caregivers who meet eligibility criteria will be provided with brief information about the study, including its timeline and expected commitment, and will be invited to participate. Those expressing willingness to participate will be provided with a detailed explanation of the study procedures, after which verbal informed consent will be obtained. Caregivers who give consent will be considered officially enrolled in the study. Following enrolment, participants will be allocated into online support groups in batches of 10 individuals.

### Intervention

The intervention will be a digitally delivered multicomponent package to enhance self-efficacy among primary caregivers of children with SMA and/or DMD. It will be developed by a team of internal experts (for the study, referred as such for their help with the designing the intervention and self efficacy scale). They include a neuromuscular specialist, paediatric neurologist, physiotherapist, occupational therapist, dietician, speech therapist and a neuroepidemiologist. The intervention will be delivered over 8 weeks in groups of 10 participants per batch. Group composition will be guided by key participant characteristics, including estimated socioeconomic status and/or the age of the care recipient/patient (e.g., <2 years, 2–7 years, >7 years), such that participants with broadly similar profiles grouped together to facilitate effective interaction and engagement during the intervention. The intervention will comprise of three core components: (i) asynchronous educational materials (video clips and pictorial guides), (ii) live sessions, and (iii) scheduled audio-based behavioural prompts. Live sessions will be conducted at predefined intervals (Week 1 and Week 5), while educational content and reminders will be delivered throughout the intervention period with decreasing frequency to promote gradual autonomy. Intervention exposure will be standardized through structured sessions (delivered using same scripts for each batch), and sequential follow-ups within participant groups.

A pictorial summary of this Intervention is given in **(Fig 2.)**:

**Fig 2.**
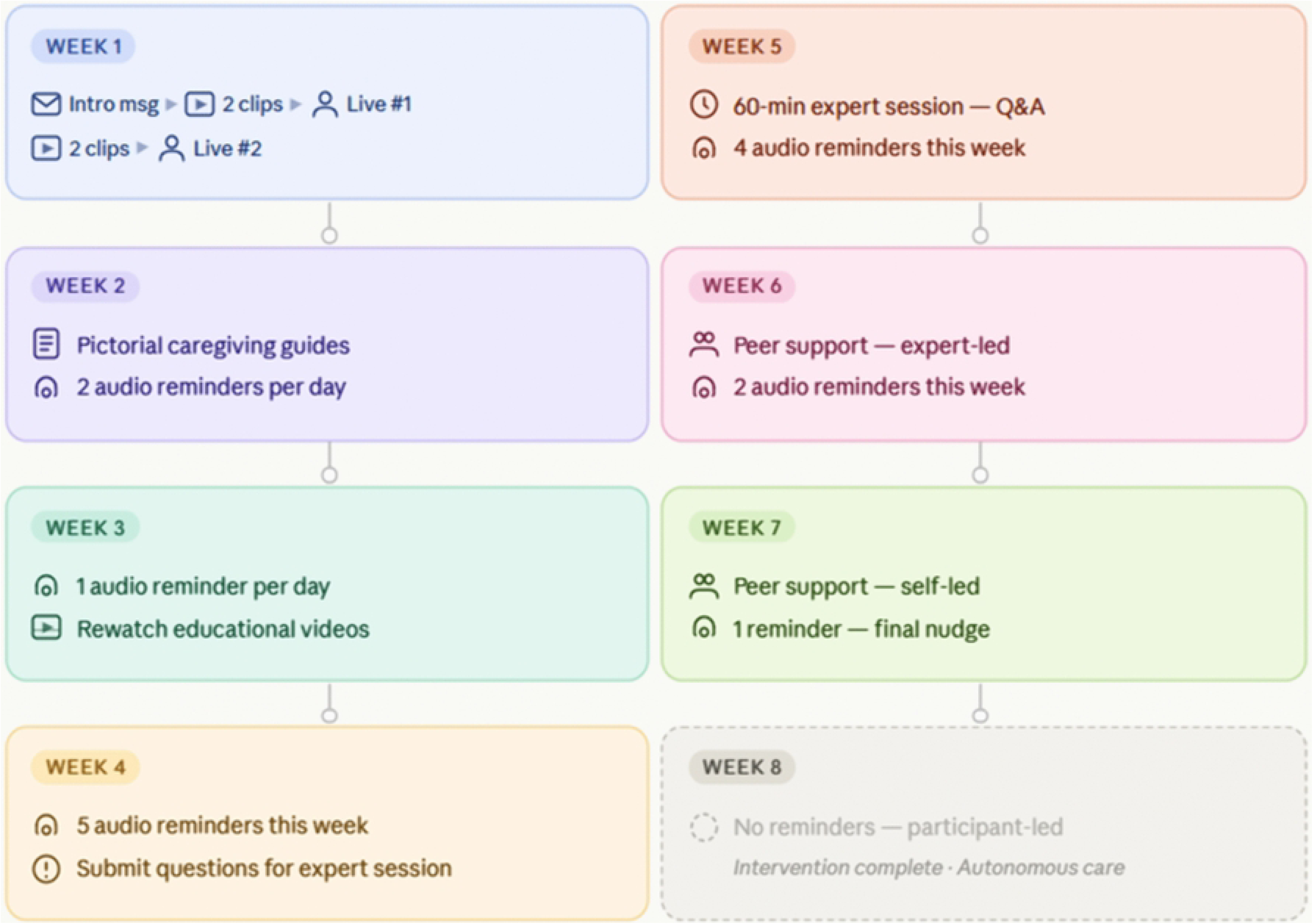
Summary of Intervention Package.

A detailed description of the Intervention, by weeks, is outlined as follows:

Week 1 ➜ At the start of Week 1, participants (caregivers) will receive an Introductory message, outlining the multi component intervention plan. This will be followed by 2 video clips, providing practical demonstrations on disease understanding and rehabilitation, on the same day. After a day’s gap, participants will attend a live session with the experts, lasting 30-35 minutes, where a group of experts will cover the same content delivered in the video clips, to reinforce their message. The following day, 2 more video clips will be shared with the participants, demonstrating solutions to further aspects of disease rehabilitation. The next day, participants will receive another live session with the experts, lasting 30-35 minutes, revising the materials shared on the video clips. Thus, this week will provide a continuous revision on foundational education on the disease and its caregiving principles.

Week 2 ➜ In Week 2, participants will receive pictorial guides covering key caregiving practices. In addition, the online platform will deliver twice-daily audio reminders, which will act as behavioural cues to facilitate the translation of knowledge and learned skills into practice.

Week 3 ➜ In Week 3, participants will receive one audio reminder daily. Prompts will encourage caregivers to rewatch previously shared video clips and apply them.

Week 4 ➜ In Week 4, participants will receive a total of five audio reminders during the week. These will encourage continued application of caregiving practices. Reminders will be delivered irrespective of prior task completion to ensure consistent exposure to behavioural cues. Participants will also be invited to submit questions or concerns to experts in preparation for the upcoming week’s session for this week.

Week 5 ➜ A second 60-minute expert-led virtual session will be conducted to address major questions related to the application of caregiving practices over the preceding four weeks. While core content will remain standardized, the session will incorporate participant queries.

Participants will also be encouraged to engage with and support one another during the session.

During Week 5, participants will receive four reminder messages, reinforcing key caregiving behaviours.

Weeks 6 ➜ The online platform will remain open for moderated peer interaction. Reminder frequency will be reduced to two audio messages per week, delivered at the beginning and end of the week, to reinforce continuity of caregiving practices while promoting increasing independence.

Week 7 à The platform will remain open for participant interaction without active moderation.

A final audio reminder will be delivered at the end of the week, encouraging sustained caregiving practices beyond the intervention period.

Week 8 ➜ In Week 8, the intervention will be stopped, and no reminders or educational materials will be delivered. This no-nudge period will allow participants to independently maintain learned behaviours without external prompts.

### Outcome

The primary outcome for analysis will be the change in total self-efficacy score between baseline (T0) and post-intervention (T1). Subscale scores (Information, Skills, Sourcing) will be analysed as secondary outcomes. The research team will undertake a structured mapping process to ensure that self-efficacy scores and the intervention is contextually appropriate and conceptually aligned with the caregiving realities of families managing Duchenne Muscular Dystrophy (DMD) and Spinal Muscular Atrophy (SMA) patients in local setting. To measure this self-efficacy, the study will adopt the Duchenne Muscular Dystrophy Caregiver Self-Efficacy Scale (DMD-CSES), a psychometrically validated instrument originally developed for caregivers of individuals with DMD in Turkey (11). The DMD-CSES is a self-administered questionnaire that assesses caregiver confidence across three core domains: Information (knowledge and understanding of the disease and caregiving requirements), Skills (ability to implement caregiving practices such as nutrition, rehabilitation, and symptom management), and Sourcing (ability to access external support systems, including healthcare services, financial resources, and time management). It consists of 18 items rated on a 5-point Likert scale ranging from strongly disagree to strongly agree, and total scores are calculated by summing all responses, with higher scores indicating higher caregiver self-efficacy. Although this scale has demonstrated strong psychometric properties in its original setting, no culturally validated version is currently available for use in Pakistan, and its applicability to both DMD and SMA caregivers within a low-resource healthcare context has not been formally established. Therefore, cultural adaptation and content validation are essential steps to ensure that the scale remains meaningful, comprehensible, and relevant in the local setting before it is used as an outcome measure.

To support this adaptation process and ensure that the intervention content appropriately targets the same domains measured by the outcome scale, input will be sought from the internal experts. They will contribute targeted feedback to refine both the scale’s wordings and the intervention content, ensuring that language is understandable for caregivers of varying literacy levels and that each item reflects realistic caregiving expectations in Pakistan. Given the differences in healthcare access, rehabilitation availability, caregiver support structures, and cultural caregiving roles between high-income settings and Pakistan, it is anticipated that some contextual modifications may be required to preserve conceptual equivalence while improving local relevance. Following their input, the adapted version of the scale will undergo translation into Urdu, the most widely understood national language among caregivers, using a structured forward and backward translation approach to ensure semantic and conceptual consistency.

Thereafter, some caregivers will provide feedback on the study’s scale and finalise it for comprehensibility and appropriateness. This feedback will be gathered through in-depth interviews, conducted either virtually or in person. Simultaneously, these interviewees will also help in modifications in the Intervention, to ensure its educational content, terminology, and delivery format is aligned with the needs, norms and feasibility. Once participant’s refinements are incorporated in the scale, it will be sent to a panel of external experts for content validation process. These experts are referred as external experts as they were not involved in the study tool’s design, initially. They will include epidemiologists, neurologists, biostatisticians, mental health professionals, and rehabilitation experts and nurses. Content validation will involve systematic ratings for each item of the scale, to assess relevance and clarity, generating quantitative evidence that the adapted items adequately represent the construct of caregiver self-efficacy in the Pakistani context. Content Validity Index (CVI) will be calculated. Establishing content validity prior to the main study is critical to ensure that the primary outcome measure accurately reflects the caregiving competencies and support needs targeted by the intervention. The summary of these steps is better illustrated in **(Fig 3.)**

**Fig 3.**
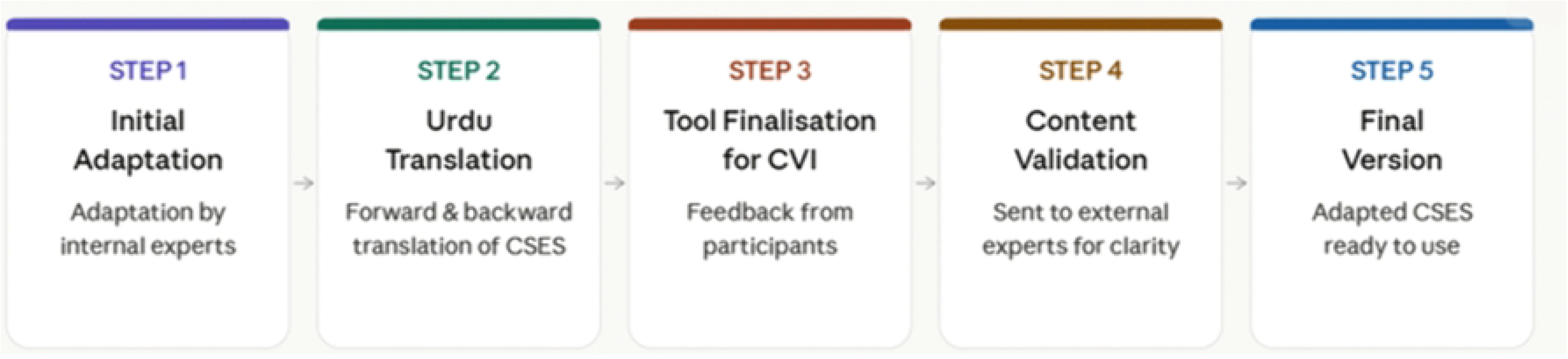
Summary of Scale Adaptation.

### Data management

All data will be collected and stored electronically and will only be accessible to authorized personnel. Since the registry has participants from all over Pakistan, the data collection would be conducted via video calls. These calls will be conducted by trained study staff. The staff will obtain a verbal consent which will be documented along with a witness. The consent will explain the study’s purpose, procedures, potential risks and benefits, and the right to withdraw from the study at any time without penalty. Participants’ identities will be protected using pseudonyms or code numbers.

During the live sessions and in unmoderated peer interactions, there is a minimal risk of heated discussion. To reduce this, participants will be given a set of rules to follow at the start of the Intervention (do’s and don’ts). Still, in case of exchange of hostile messages, the primary team members will immediately de-escalate matters. These members are medical professionals equipped with counselling experiences on handling such risks. Similarly, during live sessions the host controls will be strict (muting or disabling participants from cross-talking).

### Data analysis

Data will first be reviewed for completeness and consistency. All participants responses will be analysed. If however, data is missing (due to drop out/non-response), substitute participants will be enrolled and brought up to speed to replace if timely possible. If replacement is still not possible, they will not form a part of the final analysis and will only be reported descriptively. Cleaning procedures will include range checks, detection of outliers, and validation of logical consistency across related variables.

Descriptive statistics will be used to summarize the sociodemographic characteristics of participants, including caregiver age, gender, education level, relationship to the child, and child’s age, diagnosis, and disease duration. Continuous variables (e.g., caregiver age) will be presented using mean and standard deviation (SD) if normally distributed, or median and interquartile range (IQR) if not. Categorical variables (e.g., gender, diagnosis type) will be summarized using frequencies and percentages.

Before inferential analysis, the distribution of total self-efficacy scores and subscale scores will be assessed for normality using histograms, Q-Q plots, and the Shapiro-Wilk test. To evaluate the effect of the intervention, pre and post self-efficacy scores will be calculated. If the difference scores (pre – post) are normally distributed, a paired t-test will compare pre– and post-intervention self-efficacy scores. If normality is not met, the Wilcoxon signed-rank test will be applied.

All statistical tests will be two-sided, with significance defined as p < 0.05. Analyses will be conducted using STATA Version 17.

### Ethical considerations

Ethical approval was sought from the Ethics Review Committee of Aga Khan University (2025-11875-37040), and the trial was registered on clinicaltrials.gov by the Principal Investigator (NCT07356063), before recruitment for the study. Formal independent trial monitoring and a Data Monitoring Committee (DMC) are not required for this study. Given its exploratory nature with a small sample size and a short intervention duration, no interim analyses or stopping rules are planned. The intervention is non-pharmacological and poses minimal risk to participants; identified risks are mitigated through pre-specified strategies outlined above. Thus trial conduct, protocol adherence, data quality, and ethical compliance will be overseen directly by the principal investigators throughout the study.

All participant information will be treated as strictly confidential, and no identifying data will be disclosed in any publication or presentation. Study’s data will not be made publicly available. However, reasonable requests for anonymised data sharing for research purposes may be considered on a case-by-case basis after completion of the study and publication of the primary findings, subject to ethical approval and data protection considerations.

## Discussion

This protocol describes a single-arm, pre–post evaluation of a structured online support intervention designed to enhance caregiving self-efficacy in parents of children with SMA/DMD. The intervention is theory-based and targets key domains of caregiving confidence through education, skills reinforcement, and peer-supported engagement.

While the study addresses an important gap, several limitations warrant discussion. The relatively small sample size limits statistical power and generalisability, and findings should therefore be interpreted as exploratory. Additionally, without a comparator group, observed changes in self-efficacy cannot be attributed exclusively to the intervention, as external influences such as temporal effects or regression to the mean may contribute. Furthermore, selection bias may arise since caregivers will be recruited from a registry and must have smartphone/internet access, so the sample may over-represent more connected or resourced families. Another limitation is potential social-desirability bias (caregivers may overstate confidence), as we rely on interviewer-administered self-report measures. Despite planned grouping, based on disease-related and socioeconomic characteristics, heterogeneity in engagement, uptake, and perceived benefit may persist between groups. Variability in how participants interact with different components of the intervention may influence observed outcomes. Lastly, as the study is positioned as a preliminary effectiveness evaluation rather than a formal feasibility or implementation study, structured measures of adherence and acceptability will not be included. However, intervention exposure will be standardised through structured delivery and consistent sequencing of components across all participant groups.

Despite these limitations, the study incorporates several strengths. First, the participants will be engaged during the scale development phase, to refine both the outcome measure and intervention content, enhancing comprehensibility. Second, the intervention fidelity is bolstered by the close alignment between content development and delivery: the same multidisciplinary experts who designed the curriculum also guide the live sessions. This coherence enhances internal consistency of the intervention. Third, to our knowledge no prior study has evaluated a structured multicomponent intervention targeting caregiver self-efficacy in SMA/DMD, so this project fills a novel gap. Fourth, we will use a content-adapted outcome measure: the DMD Caregiver Self-Efficacy Scale is being carefully translated, culturally adapted and validated (content validity index, CVI, calculation) before use. Establishing the scale’s relevance and reliability in Urdu ensures that the primary outcome truly reflects local caregiving contexts. Fifth, the fully digital delivery is a major advantage in Pakistan’s context: by removing geographic barriers, the intervention can reach families nationwide at low cost. The World Health Organization encourages leveraging mobile and internet platforms to extend health education in resource-limited settings (27). Finally, embedding recruitment in the national NMD registry allowed systematic identification of eligible caregivers (an efficient approach given the rarity of SMA/DMD).

The intervention is explicitly based on social cognitive theory, which posits that self-efficacy can be enhanced through mastery experiences, vicarious learning, verbal persuasion and emotional support (28, 29). Our program provides these elements: educational videos and expert sessions build knowledge (mastery), caregiver peer interactions provide modelling (vicarious learning), and positive reinforcement cues (audio reminders) serve as persuasive encouragement. By systematically targeting the three domains of self-efficacy (Information, Skills, Sourcing) assessed by the scale, we expect incremental gains in confidence as caregivers successfully apply new practices. This theoretical linkage underscores the plausibility of the intervention improving self-efficacy.

Findings from this study will be disseminated through publication in peer-reviewed scientific journals and will also be presented at relevant conferences and professional meetings focusing on neuromuscular disorders. In addition, key results may be shared with relevant stakeholder communities, including patient support organisations and the Treat-NMD registry network, to inform future caregiver support initiatives and service planning.

Any protocol amendments will be formally documented and submitted to the Aga Khan University Ethics Review Committee for approval prior to implementation. This ensures any modifications are ethically vetted. In the unlikely event that the study must be terminated early, the principal investigator will inform all participants and the ethics committee immediately. Data collected up to termination will be analysed as feasible, with a clear statement of the decision and its impact on results. No interim changes will be made without prior ethical clearance.

This protocol outlines a theory-informed, digitally delivered intervention addressing an important gap in caregiver support for neuromuscular disorders. While the study is exploratory in nature and subject to inherent limitations, it is expected to provide valuable insights into the potential role of structured online interventions in improving caregiver self-efficacy in resource-constrained settings.

## Data Availability

No datasets were generated or analysed during the current study. All relevant data from this study will be made available upon study completion.

## Acknowledgements

Assistance was sought from Tania Furqan, Fatima Noor, Amber Amir, Ramsha Siddiqui & Dr. Khairunnissa Mukhtiar for the initial development of this study’s Intervention and Scale.

ChatGPT was used to assist with improving grammar, language fluency, and clarity of writing. Claude was used for the creation of illustrations (figures) included in the manuscript. These tools were used solely to support presentation and visualisation. All scientific content, study design, analysis plan, and interpretations were developed by the authors. The authors take full responsibility for the accuracy, integrity, and originality of the work.

## Authors’ contributions

Conceptualization: NIA

Data Curation: NIA

Formal Analysis: NIA

Methodology: NIA, WK

Project Administration: SK

Resources: SK

Supervision: SK

Validation: NIA, WK, SK

Writing – Original Draft Preparation: NIA

Writing – Review & Editing: NIA, WK, SK

## Supporting Information

S1 Checklist (SPIRIT)

S2 Checklist (TIDieR)

